# Machine learning for the prediction of urosepsis using electronic health record data

**DOI:** 10.1101/2024.05.28.24306956

**Authors:** Varuni Sarwal, Nadav Rakocz, Georgina Dominique, Jeffrey N. Chiang, A. Lenore Ackerman

## Abstract

Urosepsis, a medical condition resulting from the progression of urinary tract infection (UTI), is a leading cause of death in hospitals in the United States. Urosepsis commonly occurs due to complicated UTI and constitutes approximately 25% of all sepsis cases. Early prediction of urosepsis is critical in providing personalized care, reducing diagnostic uncertainty, and ultimately lowering mortality rates. While machine learning techniques have the potential to aid healthcare professionals in identifying potential risk factors, and high-risk patients, and recommending treatment options, no existing study has been developed so far to predict the development of urosepsis in patients with a suspected UTI presenting to an outpatient setting. In this research study, we develop and evaluate the utility of multiple machine learning models to predict the likelihood of hospital admission and urosepsis diagnosis for patients with an outpatient UTI encounter, leveraging de-identified electronic health records sourced from a large health care system encompassing a wide range of encounters spanning primary to quaternary care. Inclusion criteria included a positive diagnosis of urinary tract infection indicated by ICD-10 code N30 or N93.0 and positive bacteria result via urinalysis in an ambulatory setting (primary or emergent care settings). For these patients, we extracted demographic information, urinalysis findings, and any antibiotics prescribed for each instance of UTI. Reencounters we defined as all encounters within seven days of the initial UTI encounter. The reencounters were considered urosepsis-related if matching positive blood and urine cultures were found with a sepsis ICD-10 code of A41, R78, or R65. A variety of machine learning models were trained on this rich feature set and were evaluated on two tasks: the prediction of a reencounter leading to hospitalization, and the prediction of Urosepsis. Model performances were stratified by the patient ethnicities. Our models demonstrated high predictive performance with an area under the ROC curve (AUC) of 79.5% AUC and an area under the precision-recall curve (APR) of 13% APR for reencounters, and 90% ROC and 31% APR for Urosepsis. We computed shapley values to interpret our model predictions and found the patient age, sex, and urinary WBC count were the top three predictive features. Our study has the potential to assist clinicians in the identification of high-risk patients, making more informed decisions about antibiotic prescription and providing improved patient care.

## Introduction

Sepsis is a leading cause of death in United States hospitals, accounting for half of all hospital deaths.[1] Urosepsis, defined as sepsis progressing from urinary tract infection (UTI), comprises approximately 5-7% of all severe sepsis cases and has a mortality rate of 14% for community-acquired infections.[2] Early identification of urosepsis is crucial, as administering appropriate antibiotic treatment, providing supportive therapy, and identifying complicating factors, such as urinary obstruction, dramatically improves mortality rates.[3]Progression to urosepsis in adults is rare, however, occurring in only approximately 0.4% of all UTI.[4]

Even though only a small proportion of patients will progress to systemic infection, antibiotic treatment of suspected UTI remains common practice; physicians routinely cite this fear as a reason for prescription of antibiotics even when they are not indicated.[5] As many as 75% of antibiotics prescribed for outpatient UTI are inappropriate, including unnecessary prescriptions, excessive duration of antibiotic therapy, and misuse of broad-spectrum antimicrobials.[6–10] The self-reported annual incidence of UTI in women is 12% with a lifetime prevalence of approximately 60%,[11, 12] making the antibiotic burden attributable to these inappropriate prescriptions a significant contributor to the worsening global crisis of antimicrobial resistance. UTI are one of the most common indications for antibiotics at outpatient visits to physician offices and emergency departments, comprising almost 25% of all outpatient antibiotics prescribed.[13] Randomized placebo-controlled trials have shown, however, that antibiotic treatment for UTI offers only mildly faster symptomatic improvement compared to placebo in patients with urinary symptoms alone presenting in an outpatient setting.[14–17] Indeed, the incidence of pyelonephritis in patients presenting to office-based settings is low and is not substantially different in individuals receiving antibiotics from those treated with supportive analgesics and hydration.[18, 19]

Given the rising threat of antimicrobial resistance, improved ability to identify the small subset of patients at risk of systemic progression would improve both care and antimicrobial stewardship, but can be challenging; early signs and symptoms in patients that progress to urosepsis are similar to uncomplicated patients whose condition self-resolves without intervention. Machine learning models offer a powerful approach to evaluate patients based on their predicted mortality or morbidity and to predict required resources for more efficient management.[20] Deriving disease subtypes from patient features that are available from electronic health records (EHRs) can help clinicians make better decisions for healthcare operation policies and guide next-generation personalized medicine.[21, 22]

While numerous studies have focused on the prediction of sepsis using machine learning, few focus specifically on the progression of UTI to urosepsis. Existing research efforts have predominantly centered on specific clinical scenarios related to urosepsis, such as the differentiation of urosepsis from a urinary tract infection in inpatients,[23] predicting the risk of sepsis after flexible ureteroscopy,[24] and antibiotic prescriptions for UTIs.[25] Notably, previous investigations have not explored the domain of forecasting urosepsis risk among a group of community-dwelling individuals initially diagnosed with a UTI, the bulk of individuals receiving antibiotic prescriptions. To date, only a singular study[26] has attempted to study the association between antibiotic treatment for UTIs and severe outcomes in elderly patients in primary care settings. It is important to note, however, that this study used any hospital admission as a proxy for urosepsis, which is problematic; hospital admissions can result from a broad variety of medical conditions, of which urosepsis is a rare outcome of interest. The usage of a broad inclusion criteria can lead to incorrect inferences, especially for older patients, as hospital visits typically increase with age for numerous reasons. In fact, higher antibiotic burdens over time are associated with increased numbers of hospital admissions for a wide range of infections and antibiotic side effects.[27–29]

This study sought to apply machine learning algorithms to predict the risk of urosepsis in patients presenting with a UTI in the outpatient setting, an area ripe for harnessing the potential of machine learning to improve patient care. We sought to derive a predictive algorithm for urosepsis risk, intentionally using a minimal number of features which could be obtained easily at the outpatient point-of-care to facilitate ease-of-use and ensure applicability across a range of care delivery settings.

## Methods

### Data

De-identified electronic health records were extracted from a single academic medical health system spanning primary to quaternary care. These data were deemed non-human subjects research by the local Institutional Review Board (IRB#21-001403). Inclusion criteria included a positive diagnosis of urinary tract infection indicated by ICD-10 code N30 and N93.0 and positive bacteria result via urinalysis in an ambulatory setting (primary care, urgent care, or emergency encounter). For these patients, we extracted demographic information, urinalysis findings, and any antibiotics prescribed for each instance of UTI. These features were indexed using the patient and encounter levels to account for multiple instances. Reencounters we defined as all clinical encounters within seven days of the initial UTI encounter. The reencounters were considered urosepsis-related if matching positive blood and urine cultures were identified with a sepsis ICD-10 code of A41, R78, or R65.

### Data preprocessing

The data matrix was constructed such that each row in this dataset corresponded to a unique outpatient encounter per patient, with the columns representing the various features of an encounter. Two predictor vectors were generated, denoting whether or not within seven days of each outpatient encounter, (1) a patient was hospitalized for any cause, and/or (2) the patient was hospitalized for a UTI-related complication. The extracted features included demographics such as patient age, BMI, ethnicity (White, Latinx, Black, Asian), sex, urinalysis features such as red blood cell (RBC), white blood cell (WBC), and squamous epithelial cell numbers per unit volume, and antibiotic prescriptions, including nitrofurantoin, fosfomycin, trimethoprim/sulfamethoxazole, cephalexin, ciprofloxacin, amoxicillin, amoxicillin/clavulanate, doxycycline, and levofloxacin. Preprocessing was done to extract and map categorical variables to numerical values, perform feature engineering to extract meaningful features, remove unformatted values and outliers lying outside three standard deviations, and merge multiple antibiotics into binary columns. The features were standardized by removing the mean and scaling to unit variance, before being input to the logistic regression model. Null imputation was performed in case of missing values. Demographics were dropped before computing overall model performance. The pipeline for model development is shown in **Figure 1**.

**Figure 1:**
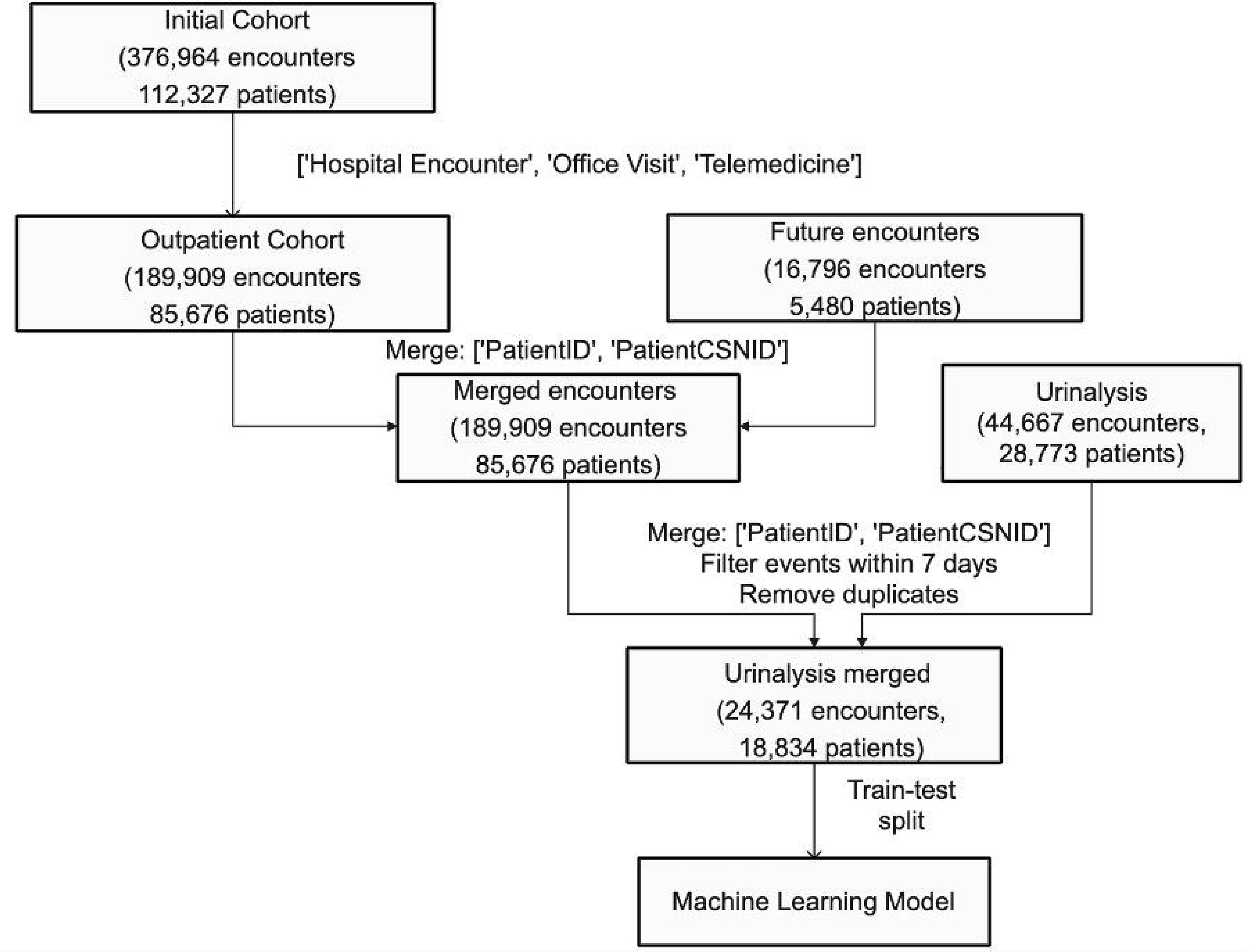
Data preprocessing pipeline for input into machine learning model to predict hostpialization or urosepsis after outpatient presentation for urinary tract infection (UTI).

### Machine learning analysis

This analysis sought to train a model that could predict the probability of an outpatient with a UTI having a reencounter due to urosepsis. We split our examination into two major analyses. First, we predicted the risk of a patient with a UTI being admitted to the hospital for any reason in a 7-day window. Next, we narrowed down our predictor variable to consider only reencounters that were demonstrated cases of urosepsis. We trained several models with different complexities and interpretability for two tasks, first, predicting all hospital reencounters, and second, predicting urosepsis. The dataset was randomly shuffled and split into training and test datasets in the ratio of 75:25. We used 3 machine learning models: logistic regression,[30] decision tree,[31]and random forest[32] implemented by the sklearn library.[33] Hyperparameter tuning for each of the models was done using a grid search over parameter space on the validation set. For random forests and decision trees, the hyperparameters set were the max depth and leaf size. The random forests were set to a max depth of 10, and decision trees were set to a max depth of 5 and a minimum leaf size of 1. We used the stochastic average gradient solver (sag) for logistic regression.

Confidence intervals were generated using bootstraping by sampling with replacement.[34] The training and test data were randomly split 10 times, and models were fit on each of the shuffled datasets. These bootstraped resamples were used to determine the 95% confidence interval. Model performance was assessed using the receiver operating characteristic curve (ROC) and precision-recall curve (PRC), showing the tradeoff between positive predictive value and sensitivity for different thresholds. We chose to use PRC since our data was heavily imbalanced; precision recall curves are known to be more informative than ROC plots while evaluating binary classifiers on imbalanced datasets[35] as they evaluate the fraction of true positives among positive predictions.

## Results

### Data and data processing

The analysis dataset of 24,371 encounters from 18,834 patients was used as the input feature matrix in the machine learning models. Among these, 563 patients (2.9%) had a hospitalization within 7 days and 118 (0.4%) of these developed urosepsis (**Table 1**). While the final cohort had a substantially higher percentage of females, male gender was positively associated with urosepsis (p<0.001).

**Table 1:** Outpatient cohort dataset summary of patients’ feature information at encounter level, including demographic information and labs related to each episode of urinary tract infection (UTI). Significant p-values are highlighted in red.

| Characteristics | Count (total) | Count (reencounter) | Count (urosepsis) | Rehospitalization |  | Urosepsis |  |
| --- | --- | --- | --- | --- | --- | --- | --- |
|  |  |  |  | Coefficient (Rehospitalization) | p-value | Coefficient (Urosepsis) | p-value |
| **Total** | 24,371 | 563 | 118 |  |  |  |  |
| **Sex** |  |  |  | **-0.32** | **9e-26** | **-0.61** | **1.2e-18** |
| Male | 3,946 (16%) | 187 (33%) | 57 (48%) |  |  |  |  |
| Female | 20,423 (84%) | 375 (66%) | 61 (52%) |  |  |  |  |
| Other/ unknown | 2 (0.008%) | 1 (0.1%) | 0 (0%) |  |  |  |  |
| **Race/Ethnicity** |  |  |  |  |  |  |  |
| **Latin** | 4214 (16.92%) | 102 (18.1%) | 31 (26.27%) | 0.11 | 0.14 | **0.02** | **0.007** |
| Asian | 2145 (8.80%) | 44 (78%) | 1 (0.84%) | 0.02 | 0.88 | -0.5 | 0.19 |
| White | 13111 (53.79%) | 305 (54.17%) | 70 (59.32%) | -0.02 | 0.72 | 0.01 | 0.57 |
| **Black** | 1626 (6.67%) | 54 (9.6%) | 3 (2.54%) | **0.12** | **0.03** | -0.47 | 0.07 |
| Other/ unknown | 3249 (13.82%) | 0 (0%) | 0 (0%) |  |  |  |  |
| **BMI** |  |  |  |  |  |  |  |
| Mean(SD) | 25.94 (6.08) | 26.7 (8.57) | 26.92 (5.4) | -0.02 | 0.20 | 0.016 | 0.91 |
| **Age** |  |  |  |  |  |  |  |
| Mean(SD) | 52.56 (22.91) | 66.26 (23.27) | 61.85 (22.02) | **0.58** | **1.22e-24** | **0.26** | **0.01** |
| **Labs**<br>Mean(SD) |  |  |  |  |  |  |  |
| **RBC (/HPF)** | 7.05 (20.51) | 13.289 (26.4) | 13.41(28.02) | **0.07** | **0.01** | -0.02 | 0.30 |
| **WBC (/HPF)** | 26.22 (41.08) | 49.42 (50) | 45.14 (53.58) | **0.33** | **2.55e-22** | **0.4** | **1.0e-07** |
| Squamous Epithelial Cells (/uL) | 15.15 (37.87) | 13.02 (36.72) | 10/28 (25.06) | 0.02 | 0.10 | -0.27 | 0.63 |
| **Antibiotics** |  |  |  |  |  |  |  |
| **Nitrofurantoin** | 0.303 | 0.09 | 0.14 | **-0.48** | **0.06** | **-0.07** | **0.05** |
| **Fosfomycin** | 0.0094 | 0.04 | 0.05 | **0.07** | **0.003** | **0.08** | **7.3e-04** |
| Trimethoprim/<br>sulfamethoxazole | 0.102 | 0.008 | 0.016 | -0.02 | 0.66 | 0.04 | 0.59 |
| Cephalexin | 0.458 | 0.54 | 0.58 | -0.02 | 0.83 | 0.09 | 0.81 |
| Ciprofloxacin | 0.196 | 0.253 | 0.19 | -0.03 | 0.59 | -0.15 | 0.79 |
| Amoxicillin | 0.019 | 0.019 | 0 | -0.06 | 0.35 | -0.35 | 0.26 |
| Amoxicillin/<br>clavulanate | 0.0139 | 0.015 | 0 | -0.06 | 0.50 | -0.35 | 0.37 |
| Doxycycline | 0.0141 | 0.008 | 0 | -0.03 | 0.57 | -0.29 | 0.30 |
| Levofloxacin | 0.0271 | 0.062 | 0.048 | -0.04 | 0.46 | 0.02 | 0.74 |

Increased urinary white and red blood cells were both associated with hospitalization, while only urinary WBC were associated with reencounters for urosepsis. The ages of both patients that were hospitalized (66.3) and patients with urosepsis (61.9) were typically older than the general population (52.6). Among individual antibiotics prescribed, only nitrofurantoin was negatively associated with urosepsis (*p*= 0.05).

### Predictive Modeling

Logistic regression, decision tree, and random forests were evaluated as potential models. We used logistic regression as a baseline model and a decision tree for model interpretability. We used random forests as more complex models that have been proven to produce good results on unstructured data. The dataset was partitioned into 75% for training and 25% for testing. ROC index, precision, and recall were used as evaluation metrics. A total of 21 features were used as input to the model, including demographic information such as age, sex, BMI, medication information, and laboratory findings (**Table 1**). We also calculated feature importance scores based on the mean decrease in Gini impurity using sklearn for decision trees and random forests to improve model interpretability (**Table 2**).

For the task of predicting reencounters, all 4 models had comparable AUC, with logistic regression exhibiting the highest APR of 0.13. We observed an improvement in model performance when we limited our predicted variable to urosepsis. Random forests were the best performing models with an AUC of 0.9, and an APR of 0.31, followed by decision trees (AUC = 0.81) (**Figure 2**). Neural networks were the lowest-performing models, with an AUC of 0.76. The most important feature in the prediction of both random forests and decision trees was the patient’s age, followed by BMI and urinalysis WBC count (**Tables 2, 3**). For both models, since our dataset is severely imbalanced, with a 0.4% prevalence of Urosepsis, the APR values are much lower than the AUC values.

**Figure 2:**
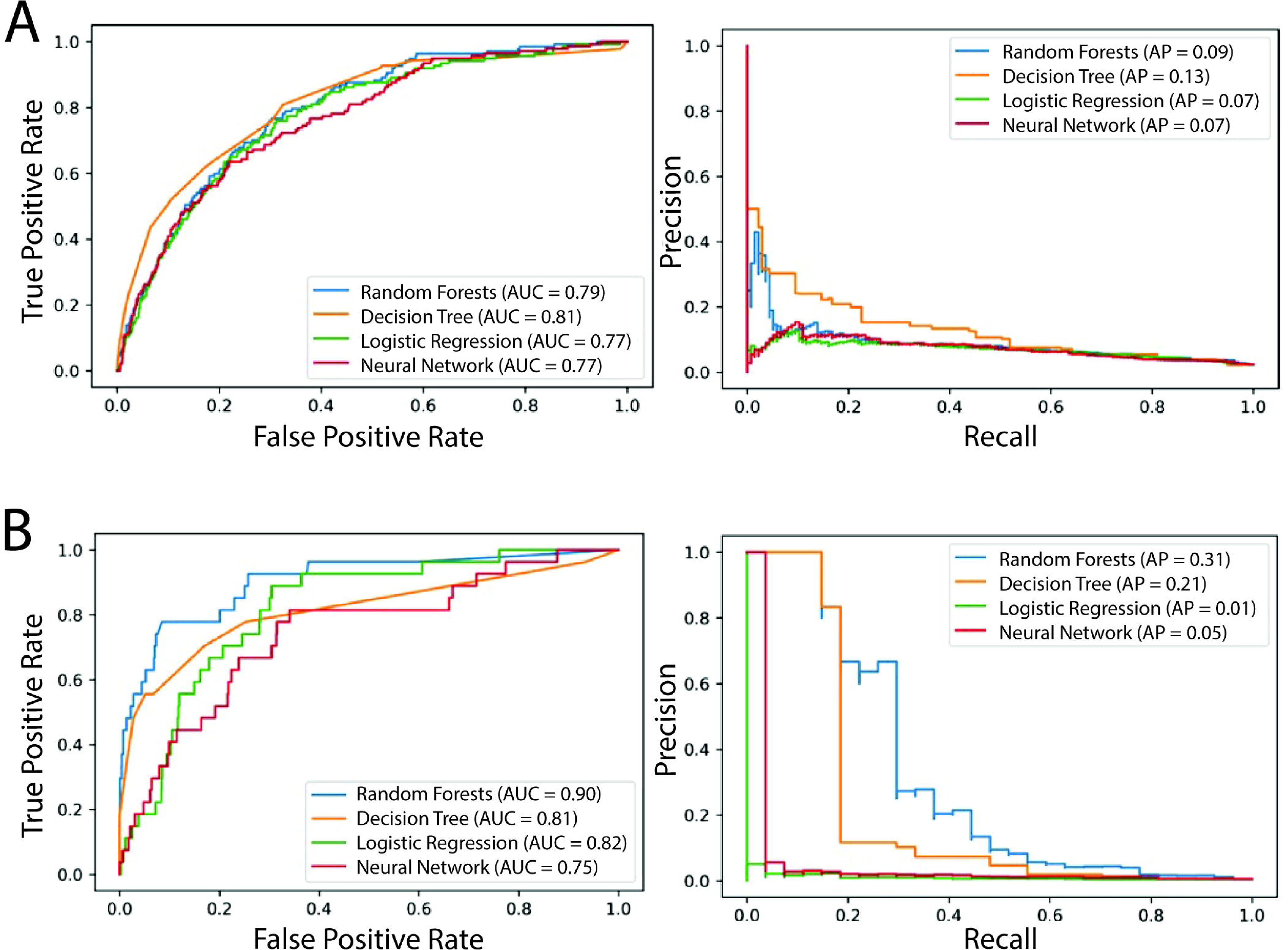
ROC and precision-recall curves for machine learning models. A) ROC and precision-recall curves for machine learning models predicting hospitalizations: for a 95% confidence interval, Decision trees achieved the highest AUC of 81%, random forests followed with 79%, logistic regression and neural networks with 77%. B) ROC and precision-recall curves for machine learning models predicting urosepsis. For a 95% confidence interval, Random Forests achieved the highest AUC of 90%, followed by Decision Trees, Logistic Regression and Neural Networks with AUCs of 81%, 82% and 75%, respectively.

### Feature Importances

To analyze the local feature importance and the directionality of the effects, we generated beeswarm plots representing the shapley values at an individual level for the random forest classifiers predicting hospitalization and urosepsis (**Fig. 3, 4**). We found patient age, sex, and urinalysis findings, including WBC and RBC counts, to be the most important features for the prediction of both hospitalizations and urosepsis. An elevated white and red blood cell count lead to a positive impact on the model output, and so did being female. We observed an interesting trend in the antibiotics, with only nitrofurantoin having a negative value on the model output when prescribed. While fosfomycin was positively associated with both hospitalizations and urosepsis, none of the other individual antibiotics prescribed demonstrated a significant association with either reencounters or urosepsis specifically.

**Figure 3:**
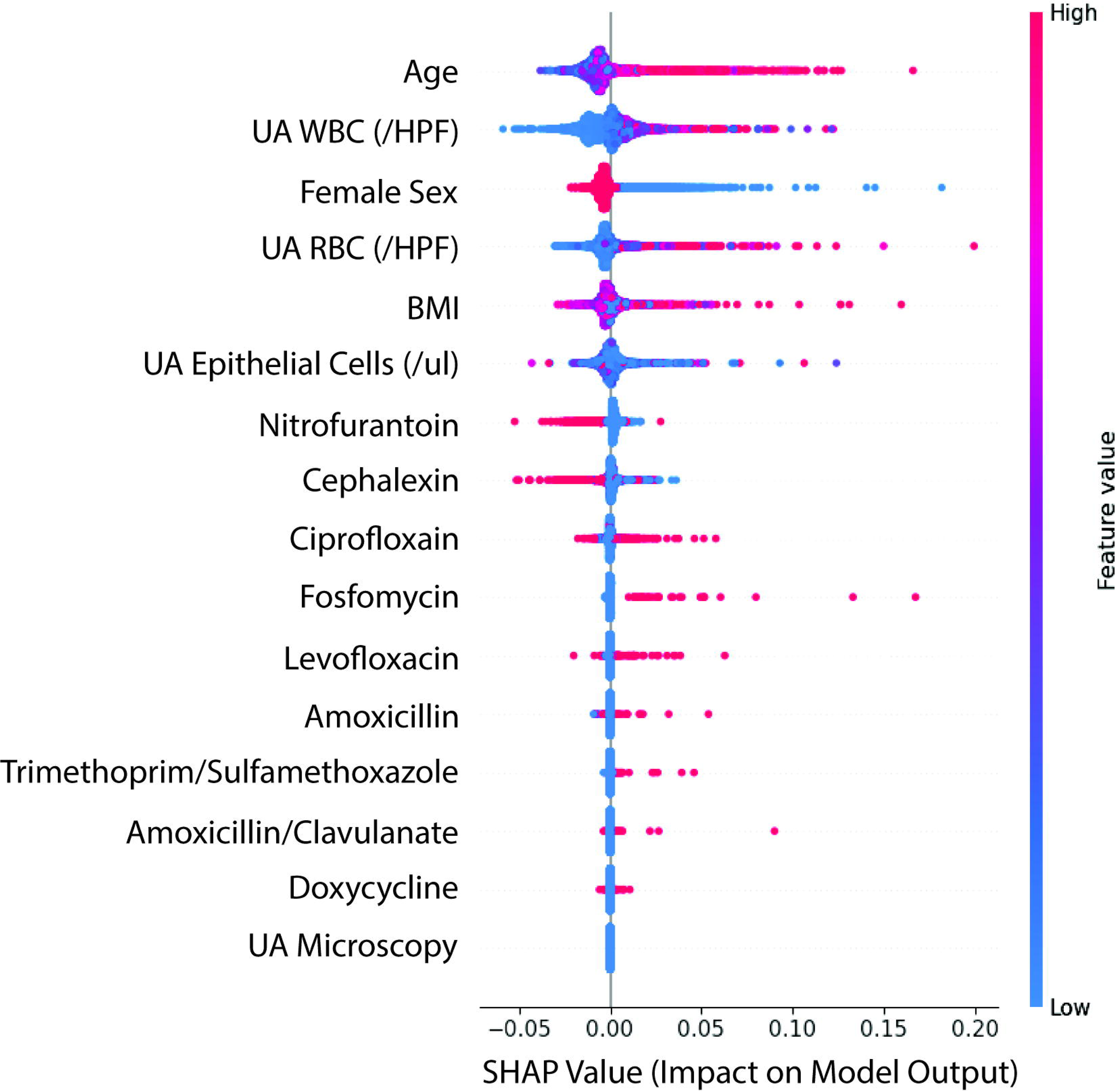
SHAP values from the random forest classifier for predicting rehospitalizations. A positive SHAP value denotes a positive impact on the model output, moving the prediction closer to +1 (no rehospitalization). A negative SHAP value denotes a negative impact on model output, moving the prediction closer to 0 (hospitalization). Each point on the plot represents an individual, and the color of the point denotes the feature value. Pink corresponds to higher values, while blue correspond to lower values.

**Figure 4:**
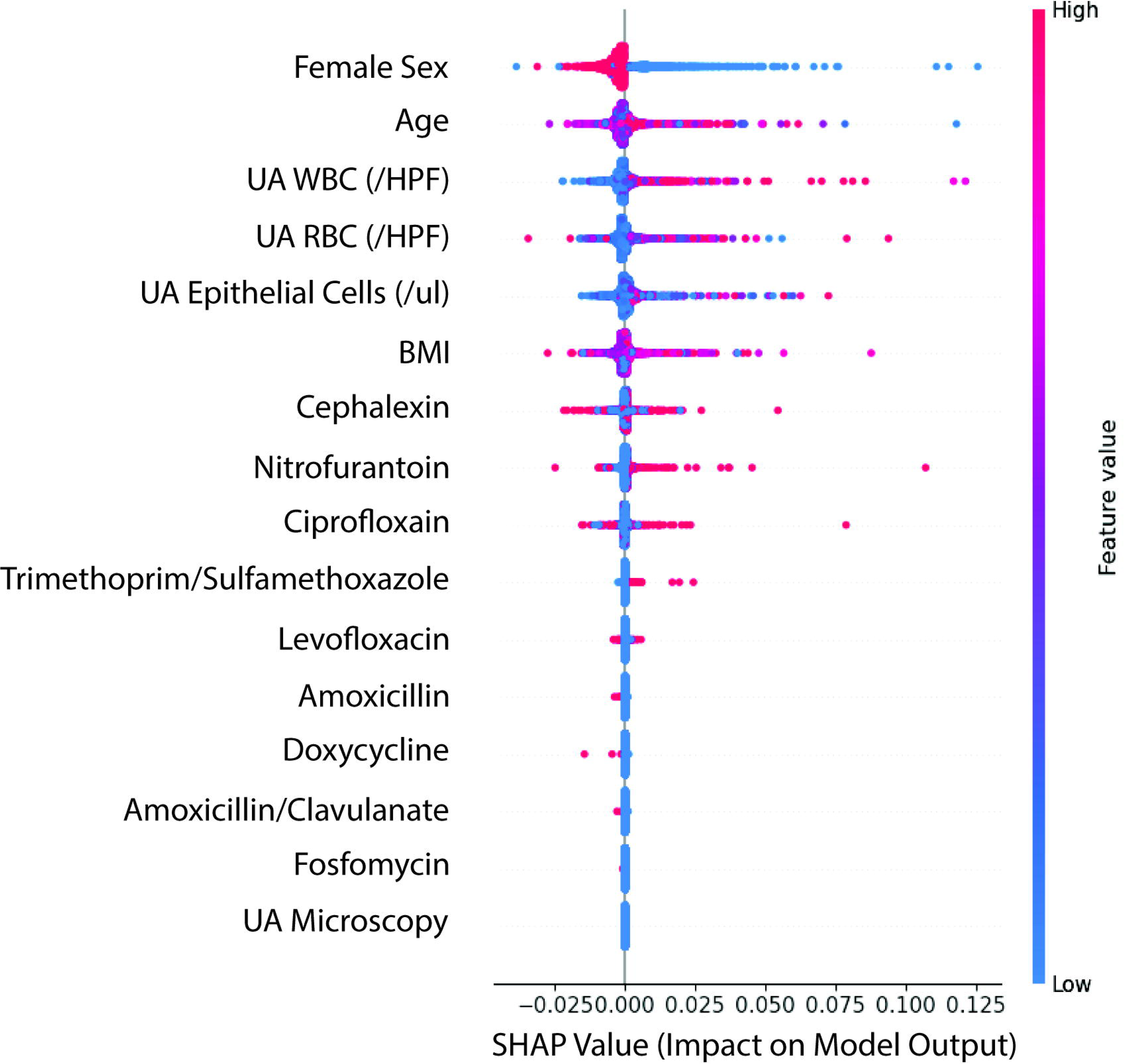
SHAP values from the random forest classifier for predicting Urosepsis. A positive SHAP value denotes a positive impact on the model output, moving the prediction closer to +1 (no urosepsis). A negative SHAP value denotes a negative impact on model output, moving the prediction closer to 0 (urosepsis). Each point on the plot represents an individual, and the color of the point denotes the feature value. Pink corresponds to higher values, while blue correspond to lower values.

## Discussion

Outpatient presentations for UTI may account for as many as 6% of all outpatient medical visits.[36] Urosepsis is a rare, but serious complication of UTI for which early recognition and antibiotic administration can be essential.[4] However, pervasive antibiotic administration for all UTI does not appreciably decrease the risk of urosepsis[14–17] and may increase poor health outcomes with increased rates of hospitalization due to antibiotic adverse events, post-infectious complications, and subsequent multidrug-resistant infections.[27–29] Thus, identification of those patients at highest risk for septic progression of UTI might allow for targeted antibiotic administration to those in whom the benefits outweigh the risks and improve antimicrobial stewardship.

In this machine learning analysis, we generate machine learning models to predict the risk of sepsis following an outpatient encounter for UTI. We first considered the primary problem of predicting all possible reencounters at the hospital, and then the narrower problem of predicting microbially-defined urosepsis. Our machine learning models had varying performances, based on model complexity and interpretability of the results, but demonstrated that random forests were the best performing models, both in terms of ROC and model interpretability. The random forest model predicted urosepsis with an excellent AUC of 0.90 using only a minimum number of patient features, all of which could be obtained at point-of-care, such as patient age, BMI, and urinary WBC counts as the top important features. While neural networks are known to handle high dimensional datasets well, in this circumstances, its performance was limited due to small dataset sizes. Logistic regression and decision tree models exhibited intermediate performance, with baseline ROC of 0.81 and 0.82, respectively.

A prime advantage of our analysis is that we have access to a unique dataset consisting of deidentified patient records spanning from primary to quaternary care, enabling us to track patient trajectories from primary care visit to hospitalization. This unique dataset, covering patients’ journeys from their primary care doctor to hospital treatment, including the pertinent demographic, vital signs, laboratory testing and microbial culture results, allowed us to build several machine learning models demonstrating excellent performance predicting the risk of hospital reencounters, both overall and those specifically caused by urosepsis, after primary outpatient presentation for UTI.

Previous analyses of urosepsis risk after outpatient presentation for UTI have primarily used claims-based data detailing any-cause hospitalization or sepsis admission as a surrogate for urosepsis[26] as there is no specific ICD-10 code for urosepsis. As bacteriuria is common in hospitalized patients, which may lead to an inappropriate diagnosis of urosepsis in patient with an alternative cause for hospitalization, we specifically examined those patients in whom blood cultures at hospitalization for sepsis reflected the same organism as that present on the urine culture obtained at the outpatient evaluation. Our data revealed that only the minority of hospital readmissions met the microbial definition of urosepsis, indicating that the majority of hospitalizations may be due to healthcare conditions unrelated to urosepsis; these data are in agreement with previous analyses demonstrating that all-cause hospitalizations were five-fold more common than urinary-infection related hospital admissions following outpatient presentation for UTI.[37]

Thus, models examining hospital readmission are limited by the accuracy of the coding of the providers involved in each encounter, which are likely flawed. Accumulating data suggests that >60% of UTI diagnoses and ∼75% of urosepsis diagnoses may be inaccurate,[38–40] which implies that reencounters should not be used as a proxy for either incompletely-treated UTI or urosepsis. Indeed, using a better-defined predictor lead to a model with higher predictive performance, and more meaningful feature importance.

Given the high frequency of bacteriuria, particularly in older and institutionalized adults, any changes in overall health, such as waxing mental status or malaise/fatigue, can be mistakenly attributed to UTI despite numerous specialty society guidelines attempting to dispel this myth.[41–43] It is possible that in the context of misdiagnosis, other conditions that may be unrecognized or untreated due to this misdiagnosis may lead to the eventual hospitalization of the patient. It is also possible that antibiotic treatment itself may contribute to hospitalization.[27–29] As many as 20% of patients prescribed antibiotics will experience an adverse effect of the medication, from allergic reactions to dehydration from antibiotic-associated diarrhea or C. difficile colitis. In addition, frequent antibiotic use is associated with increased hospital admissions for infection-related complications in a dose-dependent manner.[27] Most of these readmissions are seen in the few days following antibiotic administration, and the rates of hospital admissions for infectious complications increase with larger cumulative antibiotic exposures. [28, 29] Future studies may need to account for cumulative antibiotic burden to further improve predictive modeling.

Cumulatively, these data support the use of the more stringent model in which we defined our predictor variable narrowly by the combination of concordant blood and urine microbial cultures and a sepsis ICD-10 code of A41, R78, and/or R65. Predictive ability as measured by AUC and APR was excellent using the more precise definition of urosepsis. While a high ROC may have been high due to the increased class imbalance, the APR, which is not affected by class imbalance, was still significantly above chance. We observed several changes from the more broadly defined readmission model in the top-most predictive features, as well as feature importance scores (**Supplemental Figs. 1, 2**). While the top few important features remained the same in predicting readmissions and urosepsis, it is important to note that the importance scores for antibiotics increase for the case of urosepsis. Nitrofurantoin, widely considered the first-line antibiotic for UTI due to its good penetration into the urinary tract and minimal collateral damage, remained the only antibiotic negatively associated with urosepsis risk. Interestingly, commonly prescribed broad-spectrum antibiotics, such as the fluoroquinolones ciprofloxacin and levofloxacin, were not associated with decreased risk of urosepsis, supporting previous data demonstrating that neither increased duration nor increased broad-spectrum agents are associated with improved outcomes in UTI.[41, 42] Fosfomycin, which is also a first-line antibiotic for UTI[41, 42] and the only oral agent with efficacy against extended-spectrum β-lactamase (ESBL)-producing E. coli, was associated with increased risk of urosepsis. As fosfomycin is typically effective in UTI treatment,[44] we suspect that this association may be driven with the poor availability of fosfomycin in commercial pharmacies, frequently leading to delays in antibiotic administration. While potentially harmful when used indiscriminately, antibiotic usage plays an important role in the case of urosepsis, with delays in administration in these select cases leading to worse outcomes.

It is important to note, however, that only approximately 6% of all UTI-coded encounters had available urinalysis data for review. Of the initial dataset encompassing 377,000 visits for UTI, only 24,000 of these encounters had obtained the urinalysis data necessary for our analysis. This limitation significantly narrowed the scope of our dataset and underscores the challenges in data completeness for EHR-based studies. In addition, it highlights the need to establish expectations for the standardized evaluation of patients suspected of UTI. Most guidelines discussing the evaluation of patients with suspected UTI endorse obtaining urinary testing, typically including urinalysis and urine culture, at the time of evaluation;[41, 42] these tests can be useful in ruling out UTI and ensuring prompt and appropriate treatment when indicated.[45] While educational interventions may be needed to improve UTI diagnostic accuracy, the availability of a risk calculator utilizing this information might also enhance guideline concordance of UTI evaluation as it would provide clinicians with a more tangible interpretation for these laboratory values that could better support clinical decision-making.

Future work could improve on these results by using a larger dataset with fewer missing values, imputing missing values in the existing dataset, and performing a chart review to randomly sample patients from each category. Refined models could include additional measurements and patient features, such as prior antibiotic burden, inclusion of comorbidities and medication history, and number of prior hospitalizations, to further improve model performance. It should be noted, however, that utilization of only a minimal number of factors, all of which could be obtained at the time of evaluation, provided excellent prediction of urosepsis hospitalization, which has the potential to dramatically improve both individual consequences of antibiotic overprescribing and global antimicrobial stewardship. Our study can help the scientific and medical community identify and prioritize high-risk patients, guide treatment, and improve clinical outcomes.

## Data Availability

Data and code used in this paper are not publicly available, due to institutional data policy. The code will be deidentified and repackaged to be made publicly available if this paper is accepted for production.

---

**Figure S1:** Summary plot showing cumulative importances for the prediction of hospitalization after outpatient urinary tract infection (UTI).

**Figure S2:** Summary plot showing cumulative importances for the prediction of urosepsis after outpatient urinary tract infection (UTI).

