## Supplemental Table 1 for "Machine learning for the prediction of urosepsis using electronic health record data"

| **DECISION TREE** | | **RANDOM FOREST** | |
| --- | --- | --- | --- |
| **Feature** | **Gini Importance** | **Feature** | **Gini Importance** |
| PatientBMI | 0.246133 | PatientBMI | 0.2252 |
| PatientAge | 0.199905 | PatientAge | 0.1857 |
| UA RBC (/HPF) | 0.188479 | UA RBC (/HPF) | 0.143555 |
| UA WBC (/HPF) | 0.115509 | UA WBC (/HPF) | 0.200084 |
| UA Squamous Epithelial cells (/uL) | 0.087255 | UA Squamous Epithelial Cells (/uL) | 0.144284 |
| Sex | 0.055786 | Sex | 0.022280 |
| antibiotics | 0.040556 | antibiotics | 0.0201 |
| levofloxacin | 0.021769 | levofloxacin | 0.0071 |
| Trimethoprim/  sulfamethoxazole | 0.012397 | Trimethoprim/ sulfamethoxazole | 0.0022 |
| fosfomycin | 0.011865 | ciprofloxacin | 0.0123 |
| Amoxicillin/clavulanate | 0.010839 | nitrofurantoin | 0.0075 |
| nitrofurantoin | 0.009507 | fosfomycin | 0.0053 |
| amoxicillin | 0 | amoxicillin | 0.002595 |
| ciprofloxacin | 0 | Amoxicillin/clavulanate | 0.0034 |
| doxycycline | 0 | doxycycline | 0.0019 |
| cephalexin | 0 | cephalexin | 0.0159 |

**Table S1:** Features with their corresponding Gini feature importance scores for decision trees and random forests predicting hospitalizations after outpatient urinary tract infection (UTI).
