## Supplemental Table 2 for "Machine learning for the prediction of urosepsis using electronic health record data"

| **DECISION TREE** | | **RANDOM FOREST** | |
| --- | --- | --- | --- |
| **Feature** | **Gini importance** | **Feature** | **Gini importance** |
| PatientAge | 0.486439 | PatientBMI | 0.218145 |
| PatientBMI | 0.258316 | WBC (/HPF) | 0.191065 |
| White | 0.137243 | PatientAge | 0.181527 |
| UA_SquamousEpi (/uL) | 0.031514 | RBC (/HPF) | 0.129237 |
| Sex | 0.028404 | UA_SquamousEpi (/uL) | 0.118114 |
| RBC (/HPF) | 0.028205 | White | 0.027839 |
| WBC (/HPF) | 0.011227 | Sex | 0.023557 |
| fosfomycin | 0.007993 | Latinx | 0.021294 |
| nitrofurantoin | 0.005938 | antibiotics | 0.017326 |
| Latinx | 0.004722 | cephalexin | 0.016978 |
| Black | 0 | ciprofloxacin | 0.011022 |
| UA_MICRO | 0 | nitrofurantoin | 0.010762 |
| levofloxacin | 0 | Black | 0.009584 |
| Asian | 0 | fosfomycin | 0.008412 |
| doxycycline | 0 | levofloxacin | 0.006048 |
| amoxicillin_clavulanate | 0 | Asian | 0.004625 |
| amoxicillin | 0 | trimethoprim_sulfamethoxazol | 0.003723 |
| ciprofloxacin | 0 | amoxicillin_clavulanate | 0.000334 |
| cephalexin | 0 | amoxicillin | 0.000206 |
| trimethoprim_sulfamethoxazol | 0 | doxycycline | 0.000199 |

**Table S2:** Features with their corresponding Gini feature importance scores for decision trees and random forests predicting urosepsis after outpatient urinary tract infection (UTI).
