## Supplementary figures and images for "Machine learning for the prediction of urosepsis using electronic health record data"

### Supplemental Figure 1

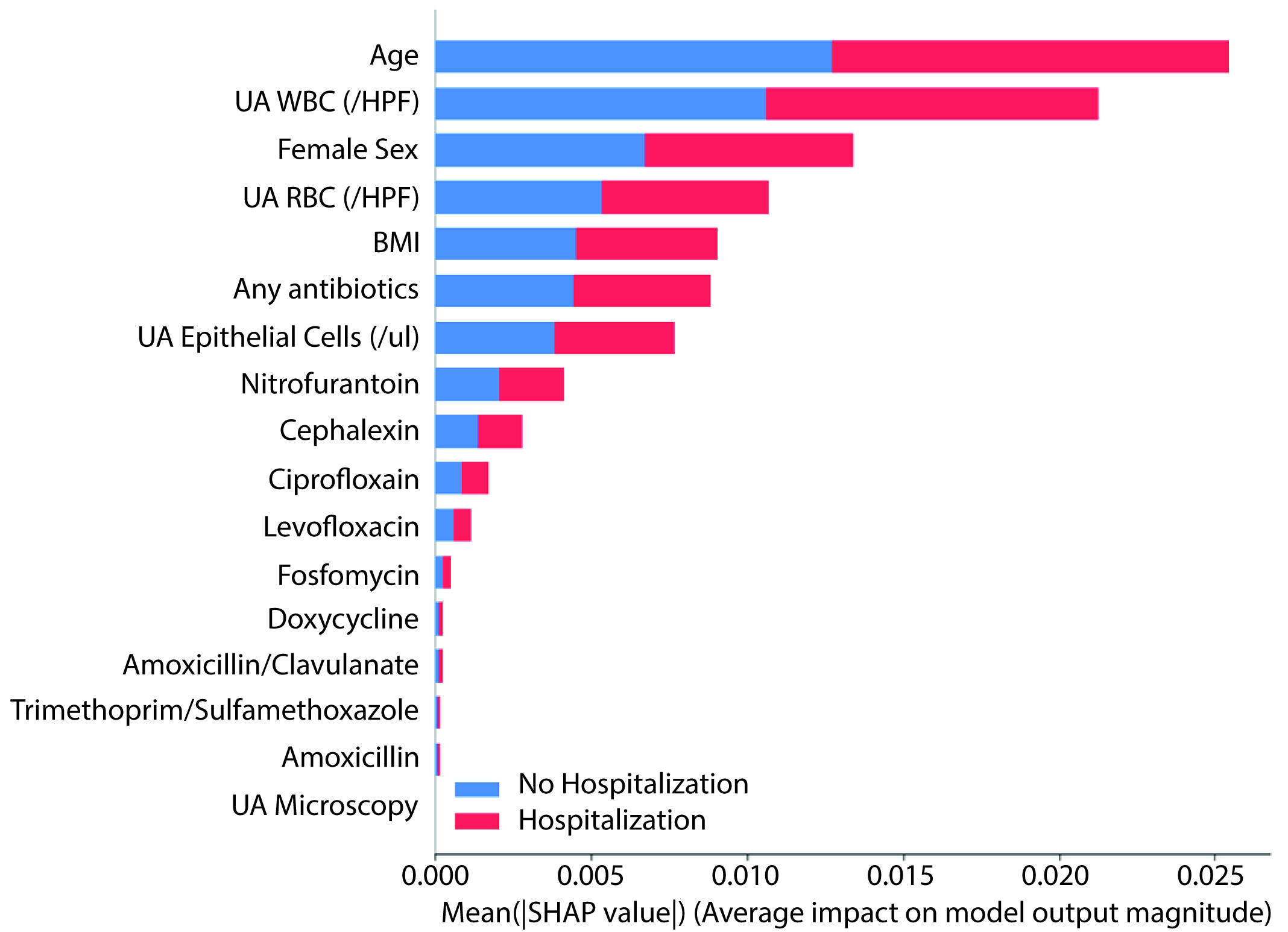

### Supplemental Figure 2

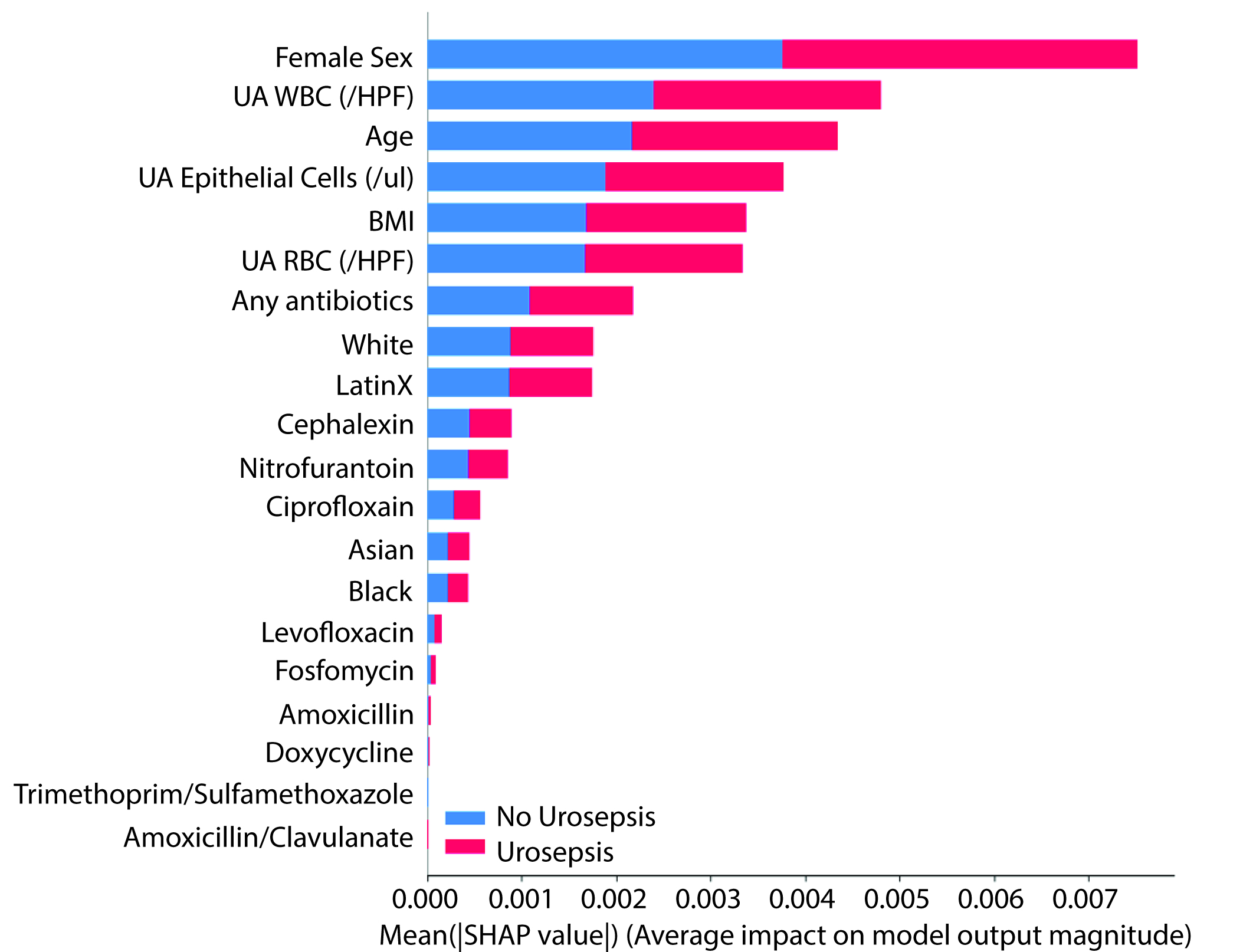
